# A qualitative exploration of pharmacists and customers barriers and facilitators to community pharmacy PrEP delivery using the COM-B model of behaviour change

**DOI:** 10.1101/2024.07.29.24311164

**Authors:** China Harrison, Hannah Family, Joanna Kesten, Sarah Denford, Jennifer Scott, Caroline A Sabin, Joanne Copping, Lindsey Harryman, Sarah Cochrane, John Saunders, Ross Hamilton-Shaw, Jeremy Horwood

## Abstract

**Objectives:** Expanding delivery of oral Pre-exposure Prophylaxis (PrEP) to community pharmacies could improve access, aligning well with government goals for England to eliminate new HIV acquisitions by 2030. Using the Capability, Opportunity, Motivation, Behaviour (COM-B) model for behaviour change, the aim of this research was to explore the barriers and facilitators of community pharmacy PrEP delivery, for pharmacists and community members.

**Methods:** Community members at elevated risk of acquiring HIV and community pharmacists were recruited to participate in semi-structured interviews. Interviews were recorded, transcribed, and thematically analysed within the framework of the COM-B model.

**Results:** 17 interviews with pharmacists (pharmacy owners n=7; employed pharmacists n=6; locums n=4) and 24 with community members (Black African women n=6; other women n=2; young adults aged 18-25 years n=6; transgender people n=6; female sex workers n=4) were carried out. Capability barriers included sub-optimal awareness and knowledge of PrEP, pharmacy facilities, and pharmacist roles in delivering public health services. Opportunity barriers included lack of staff capacity, privacy and pharmacy screening and monitoring facilities. Motivational barriers included a concern that increased access could increase sexually transmitted infections and involve a financial cost. Capability facilitators included awareness raising, HIV and PrEP training and education. Opportunity facilitators included PrEP appointments and the accessibility of pharmacies. Motivational facilitators included a preference for pharmacy delivery over other models (e.g., sexual health, GP), and a belief that it would be discrete and less stigmatising.

**Conclusion:** Pharmacy PrEP delivery is acceptable but for it to be feasible, results point to the need for the development of a behaviour change intervention focusing on education, training and awareness raising, targeting pharmacists and community members to stimulate patient activation and de-stigmatise HIV. This intervention would need to be facilitated by system and environmental changes (e.g., commissioning service).

**Key messages:** *What is already known on this topic:* Location, sigma of sexual health clinics and lack of PrEP awareness limits PrEP access for key groups among whom new HIV acquisitions remain high. Previous research and the UK government suggests PrEP provision via community pharmacies as a potential way of improving PrEP access and health equity.

*What this study adds:* This is the first research study to explore the barriers to and facilitators of pharmacy PrEP delivery for pharmacists and community members in the UK. To increase capabilities and motivation, training and awareness raising is needed. To increase opportunities and motivation, environmental and system level changes are needed.

*How this study might affect research, practice or policy:* Results point to the acceptability of pharmacy PrEP delivery, but for it to be feasible behaviour change interventions supported by system and environmental changes are needed.

## Introduction

Oral Pre-exposure prophylaxis (PrEP) became available free of charge via National Health Services (NHS) specialist sexual health clinics across England in 2020 after the Impact Trial (1). PrEP is almost 100% effective at preventing HIV acquisition when taken as prescribed (2–4). Based on its effectiveness, PrEP use has been encouraged for populations at risk of acquiring HIV and has been highlighted as integral to the UK Government’s commitment for England to eliminate new HIV acquisitions by 2030.

To initiate PrEP, eligible people require baseline and regular in clinic appointments to ensure client safety. These include screening for sexually transmitted infections (STI), HIV and kidney function tests (5). Consequently, currently in the UK PrEP is primarily accessed via face-to-face clinic-based services.

Clinic data and research exploring PrEP uptake in the UK shows most PrEP users to be white gay, bisexual, and other men who have sex with men (GBMSM) (6–8). While PrEP use among GBMSM has contributed to decreasing the number of new HIV acquisitions in the UK (7), the sexual health clinic based national PrEP delivery model could be limiting access for key groups among whom new cases of HIV remain high and PrEP use low (9). For example, in 2022, 28.4% of new HIV diagnoses were among people of black African heritage (64.1% were black African women), heterosexual men (16.8%) and women (23%). For the first time in over a decade, the number of new HIV diagnoses among heterosexuals in Scotland were higher than for GBMSM (10) and the number of women diagnosed late in England in 2022 was the highest observed since 2018 (n=239). These findings suggest the current model of PrEP provision may be restricting access for some communities, contributing to health inequalities (11). Individual and system-level barriers including the location of sexual health clinics, ability to access appointments, and stigma of sexual health clinics have been suggested to be inhibiting PrEP initiation and continuation (12). This points to the need to improve and widen PrEP access, to individuals at risk of HIV acquisition but underserved by current delivery model.

The UK government and previous research have pointed to PrEP provision via community pharmacies as a potential, effective way of improving access and health equity (6, 13–15). Pharmacists are well-positioned to support PrEP delivery due to their accessibility, medication expertise and increasing roles in providing sexual and reproductive health services to the community (16–18). Internationally, community pharmacy PrEP delivery has been associated with improvements in both PrEP initiation and continuation (19–21). Despite this success, our previous scoping review highlights several barriers to community pharmacy PrEP delivery for pharmacists and pharmacy clients (22). However, most research on these barriers has been conducted in the USA, leaving the acceptability and feasibility of community pharmacy PrEP delivery in the UK unexplored. Additionally, current UK PrEP delivery is a complex intervention, dependent on multiple stakeholders and unique commissioning pathways. Consequently, a UK community pharmacy PrEP delivery model would require behaviour change at the pharmacy, client and potentially stakeholder and commissioning levels.

The Capability, Opportunity, Motivation, Behaviour (COM-B) model of behaviour change (23) has been widely used to explore the barriers and facilitators to healthcare change interventions. COM-B posits that a behaviour (B) e.g. PrEP delivery, is influenced by an individual having i) the capabilities (C) [physically (skills) and psychologically (knowledge)], ii) opportunity (O) [social (societal influence) and physical (environmental resources)] and iii) motivation (M)[automatic (emotion) and reflective (beliefs, intentions)]. Therefore, informed by the COM-B model, we aimed to explore the perceived behavioural barriers and facilitators of community pharmacy (hereafter pharmacies) PrEP delivery for pharmacists and community members who may be at elevated risk of acquiring HIV.

## Materials and methods

### Participants and sampling

Community Pharmacy Avon (CP Avon) recruited community pharmacists in the Bristol, North Somerset, South Gloucestershire (BNSSG) area. Community members were recruited via adverts through social media and in community settings (e.g., community centres, universities) targeting individuals/communities at elevated risk of acquiring HIV. These included non-black African cis gender women, Black African cis gender women, transgender people, young people aged 18-25 years and female street sex workers. Purposeful sampling ensured diverse participant representation (e.g., gender, ethnicity).

All participants interested in participating in the study were contacted with more information and invited to participate in an interview. Participants provided written or verbal informed consent and were reimbursed for their time. Interviews were conducted between July and November 2023.

Ethical approval (12833) for the study was awarded by the University of Bristol’s Faculty of Health Sciences Research Ethics committee.

### Semi-structured interviews

Interviews were carried out face-to-face, via phone or online using Zoom or MS Teams. Interview topic guides were informed by COM-B and our aim to identify the barriers and facilitators of pharmacy PrEP delivery and were reviewed by two public contributors from our target population to ensure question appropriateness.

### Data management and analysis

Interviews were audio recorded, transcribed, imported into NVivo software and analysed using thematic analysis (24) within the framework of the COM-B. Initially familiarisation with the data was undertaken by listening to and reviewing each interview transcript. Then, in an iterative process, data was coded line by line, with all data relevant to each code collated. Codes were then deductively allocated to the appropriate domain of the COM-B thematic framework, where they were deemed to conceptually fit. Community member and pharmacist transcripts were analysed separately before being triangulated to compare similarities and differences. Data was coded and aligned to the COM-B by the first author (CH) and reviewed by the second (HF). Any disagreement was resolved through discussion.

## Results

### Recruitment outcome

Seventeen interviews with community pharmacists and 24 with community members, each lasting on average 40 minutes, were carried out. Pharmacists were pharmacy owners (n=7), employed pharmacists (n=6) and locums (n=4). The socio-demographic characteristics of the community members are presented in Table 1. Age ranged from 19-48 years, most identified as female, were heterosexual and had never used PrEP.

**Table 1.**
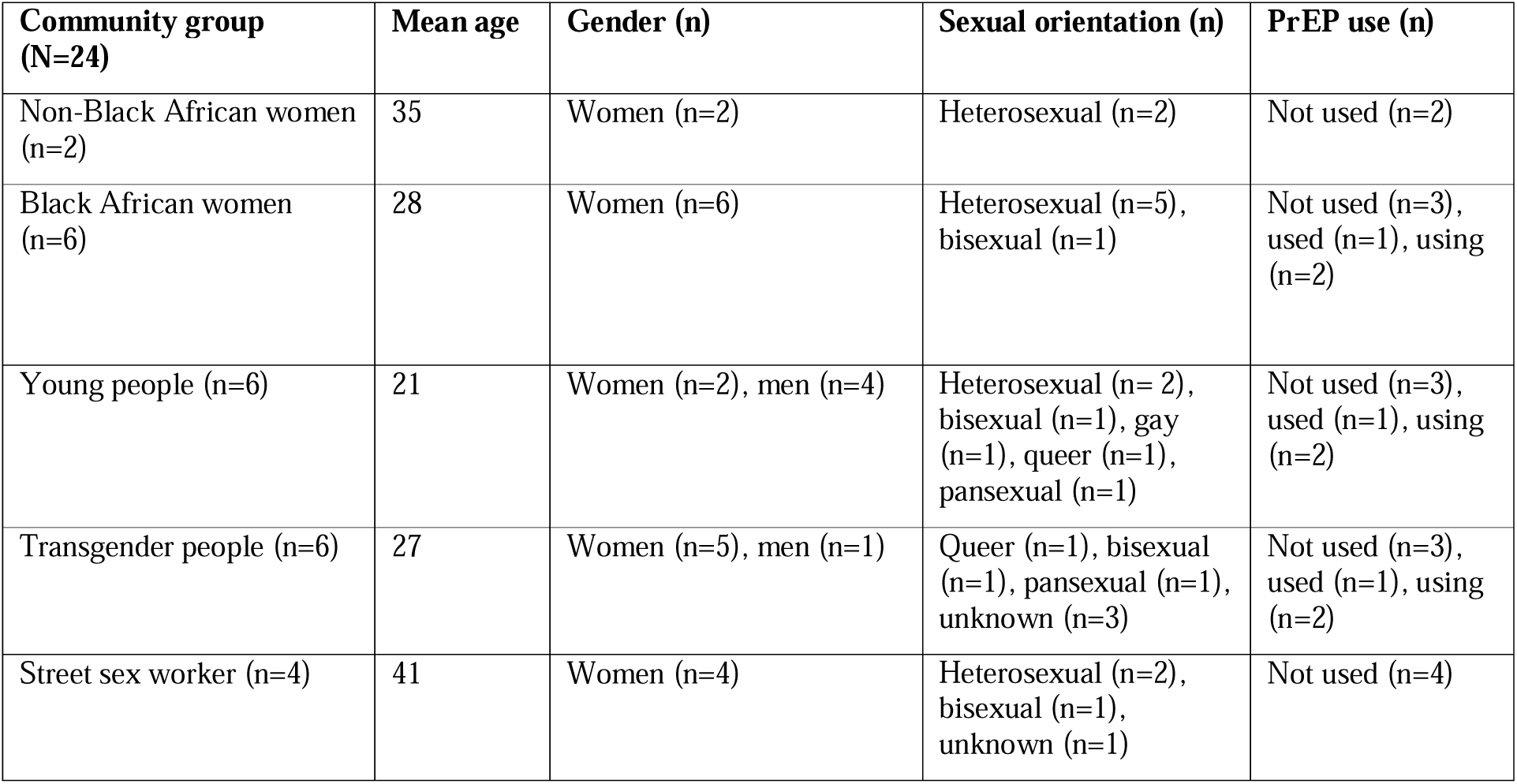
Socio-demographic characteristics of community members group.

### Barriers of and facilitators to community pharmacy PrEP delivery

The barriers and facilitators identified are presented in Figures 1 and 2 respectively and synthesised narratively below according to the COM-B with illustrative quotes in Table 2 (see supplementary Tables 1 and 2 for further illustrative quotes). Similarities and differences between the participant groups are narratively highlighted and shown using group identifiers (pharmacist (pharm), non-Black African woman (W), Black African woman (BAW), transgender person (TP), young person (YP), sex worker (SW)) followed by participant numbers. Within illustrative quotations the use of […] indicates part of the quotation was not presented because it was not relevant, whereas (text) indicates additional text added for clarity.

**Figure 1:**
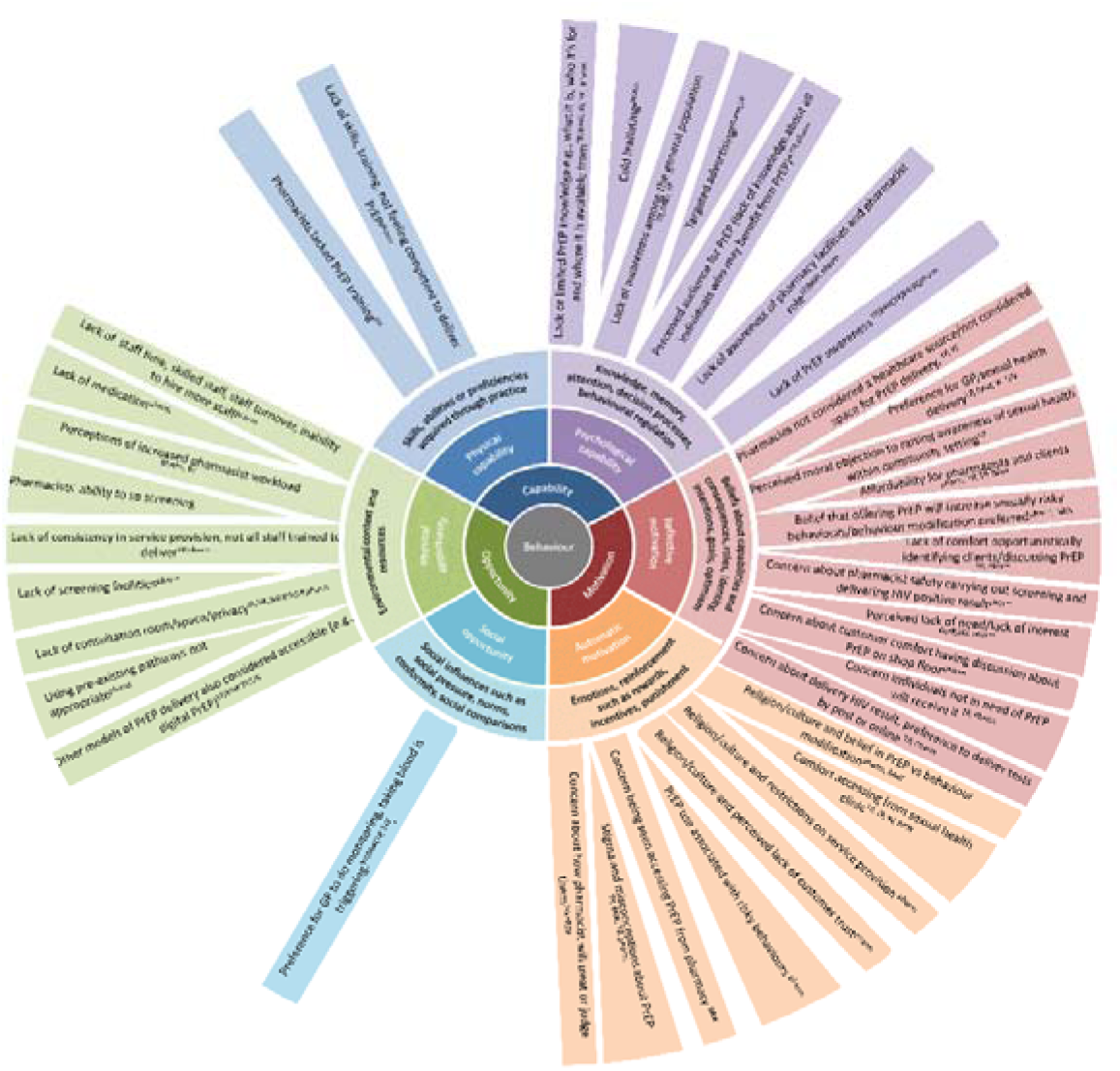
Barriers of community pharmacy PrEP delivery identified from the interviews with pharmacists and community members presented according to COM-B. (pharmacist (pharm), non-Black African woman (W), Black African woman (BAW), transgender person (TP), young person (YP), sex worker (SW)) and participant numbers

**Figure 2:**
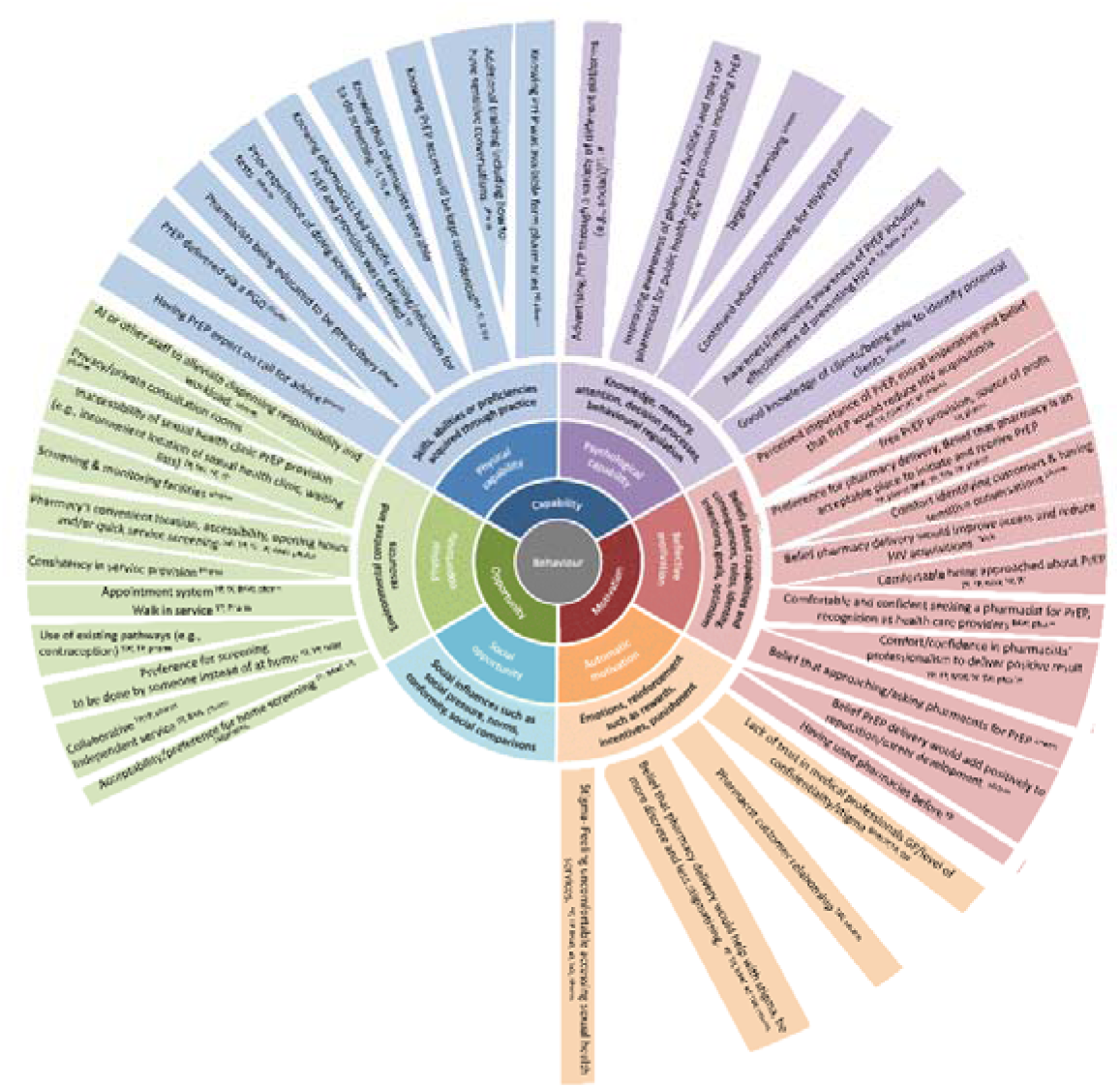
Facilitators to community pharmacy PrEP delivery identified from the interviews with pharmacists and community members presented according to COM-B (pharmacist (pharm), non-Black African woman (W), Black African woman (BAW), transgender person (TP), young person (YP), sex worker (SW)) and participant numbers.

**Table 2.**
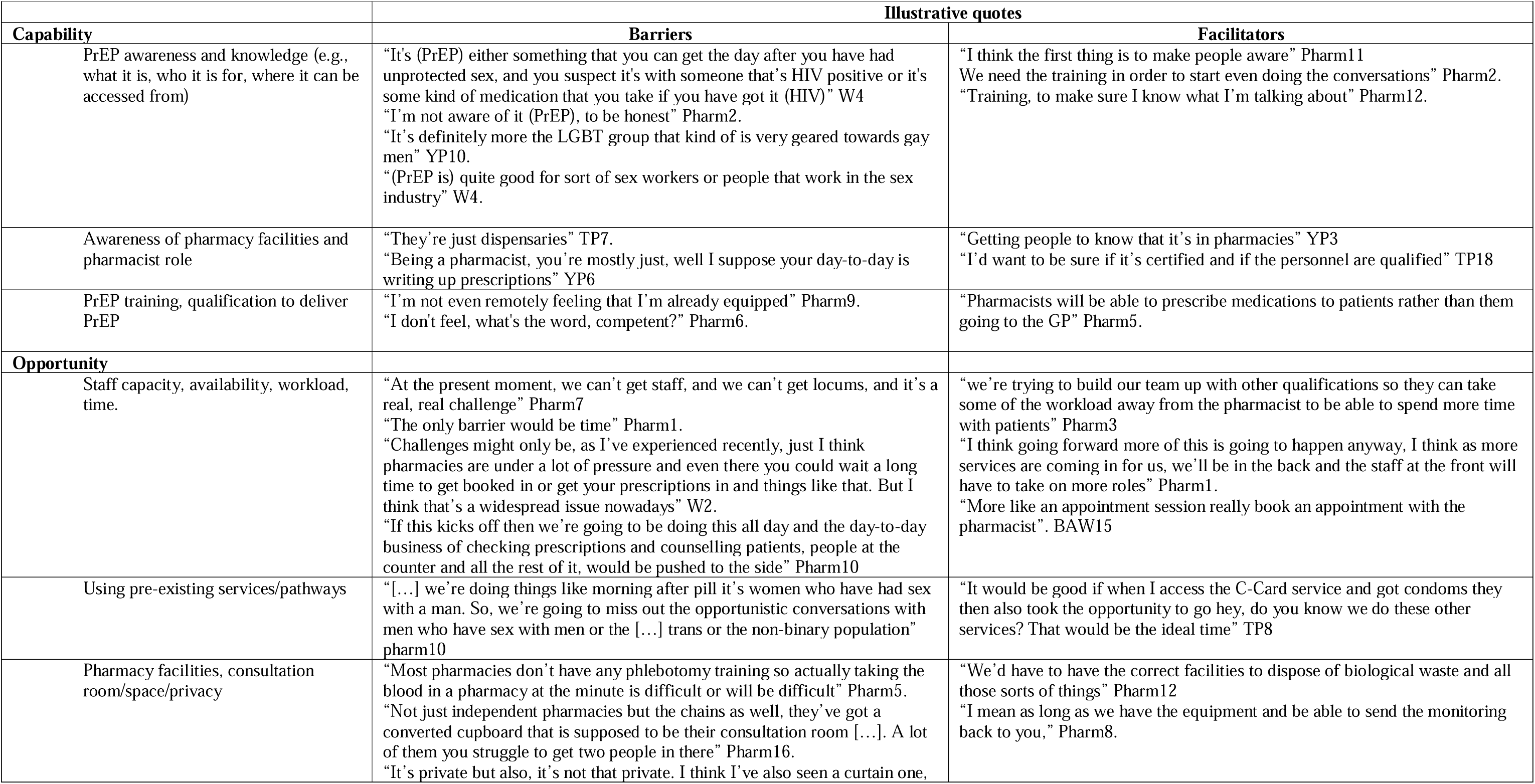
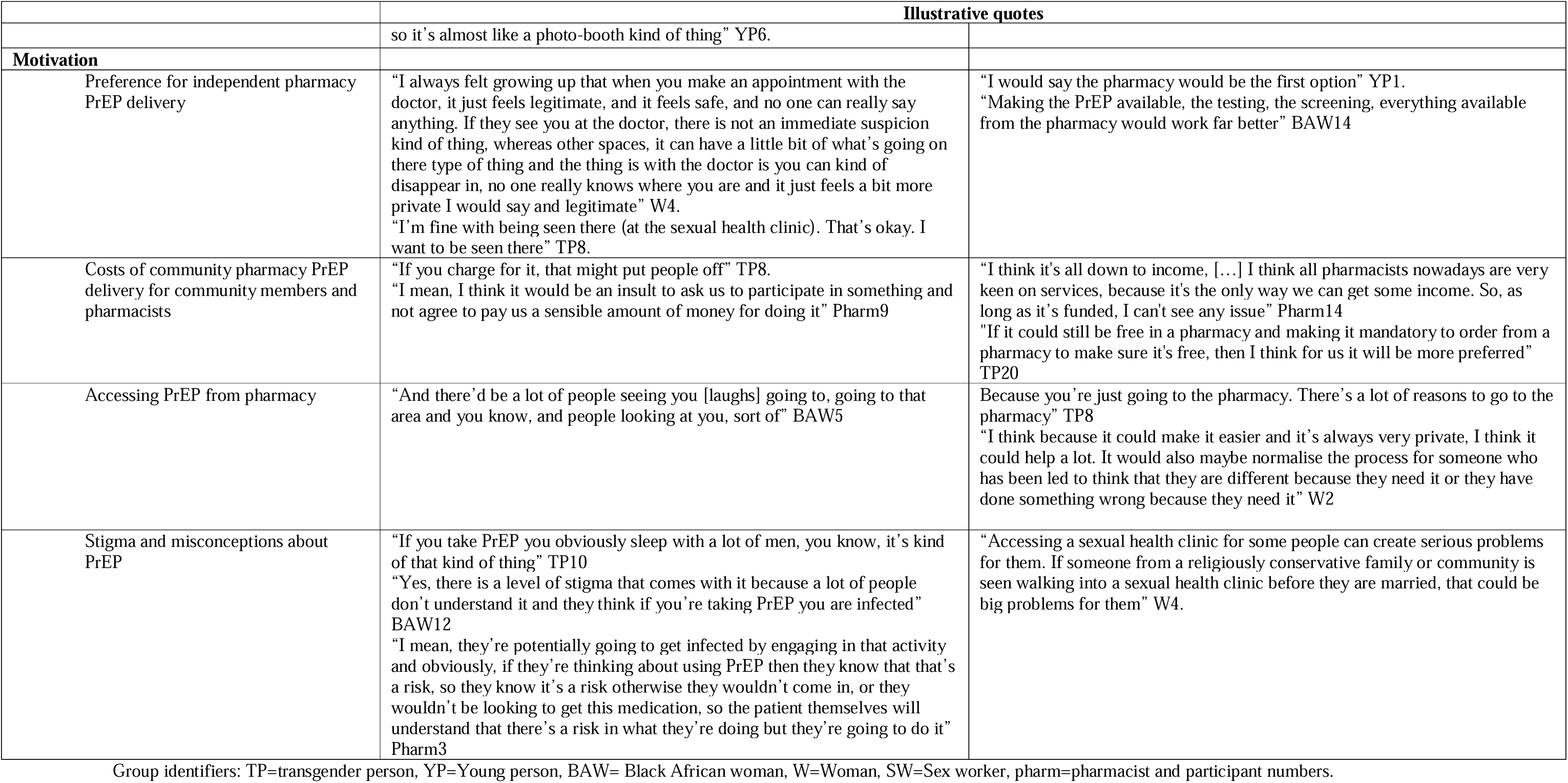
Illustrative quotes for some of the barriers and facilitators identified according to COM-B.

### Capability Barriers

Capability barriers to pharmacy PrEP delivery for community members and pharmacists included sub-optimal knowledge and awareness of PrEP (i.e., its purpose, beneficiaries and availability) with some community members and most pharmacists unaware of PrEP prior to the research and associating its use with GBMSM. Consequently, pharmacists felt unequipped with the necessary knowledge or training to deliver PrEP.

Additional barriers among community members were a lack of knowledge about pharmacy facilities (e.g., consultation rooms) and the roles, qualifications, and responsibilities of pharmacists in terms of public health service provision. Community members tended to associate pharmacies with access to over-the-counter medication and/or dispensing prescriptions rather than a point of contact for consultations for different health services, including sexual health.

### Capability facilitators

To facilitate PrEP delivery, community members and pharmacists highlighted the need to improve PrEP awareness and knowledge among pharmacists and the public through education, training, and advertisements. For pharmacists, education and training was perceived to enhance their capability and confidence to deliver PrEP. Similarly, having previous experience of carrying out similar screening tests to those that would be required for PrEP delivery (e.g., swabs, finger prick blood tests) was perceived to improve the feasibility of pharmacy PrEP delivery. For community members, particularly those from minoritized communities, knowing pharmacy PrEP delivery would be kept confidential and separate to GP records was also reported to be facilitative.

### Opportunity barriers

Opportunity barriers for pharmacists included lack of staff, high turnover, inability to hire more staff, perceived added burden of PrEP delivery to workload, and lack of physical space and privacy in pharmacies. Pharmacists and community members also highlighted barriers such as having to ask for PrEP on the shop floor in front of other customers, and the perceived lack of privacy offered by consultation rooms due to location, size, and appearance. The potential inconsistent service provision between pharmacists and pharmacies was also noted as a barrier to improving access and sustainability of pharmacy PrEP delivery. Similarly, pharmacists and community members highlighted that offering PrEP via already established sexual health services (e.g., morning after pill) within pharmacies, could restrict access to communities or demographics among whom these services are mainly sought e.g., young women.

Additional barriers reported by some community members included a preference for an experienced GP or nurse, rather than a pharmacist, to carry out the screening and monitoring tests This was particularly evident for individuals for whom having blood taken was reported to be triggering due to discomfort with needles or previous drug use. Notwithstanding this, some community members expressed a preference for digital PrEP delivery with pharmacy-based screening and monitoring because they perceived the process and results to be potentially quicker and more reliable than home testing and monitoring.

### Opportunity facilitators

Opportunity facilitators for community members and pharmacists include pharmacies’ accessibility, convenience, proximity, and walk-in service provision. This was particularly beneficial for community members with complex lifestyles who couldn’t travel to sexual health clinics, individuals living in rural areas or those seeking PrEP ‘on demand’ at short notice and potentially outside of clinic opening hours. Having a pharmacy appointment for PrEP was seen as advantageous, offering not only convenience but also privacy, avoiding the need to request PrEP publicly on the shop floor.

Although PrEP delivery via already established pharmacy sexual health services (e.g., c-card, emergency contraception) was reported to restrict access for some demographics, it provides opportunities for pharmacists to raise awareness of PrEP and signpost community members to sexual health clinics. Pharmacists and community members suggested home STI and HIV test kits prior to collecting PrEP could facilitate pharmacy dispensing. However, there were differing opinions on completing tests at the pharmacy, with some community members favouring being able to complete all tests at the pharmacy.

Pharmacist specific facilitators included having screening and monitoring facilities, the use of Artificial Intelligence Patient Medical Record (PMR) system to assist pharmacists manage and dispense prescriptions, alleviate dispensing responsibilities and free up pharmacists time, and training other pharmacy team members to take on some of the pharmacist’s workload. Community member specific facilitators included the inconvenient location of sexual health clinics and pharmacy PrEP delivery offering easier access with less bureaucracy.

### Motivation barriers

Motivational barriers for pharmacists and community members included perceiving a lack of need for PrEP, personally or within the community, concern about unnecessary use by individuals not in need, and the potential financial costs associated with pharmacy PrEP delivery. Personal and systemic stigma was also seen as a potential barrier particularly among sex workers, ethnic minorities and transgender communities who have faced stigmatising interactions in healthcare settings before.

Pharmacist-specific barriers included concerns that increasing PrEP access could lead to riskier sexual behaviours and more STIs, a preference for promoting behaviour modification over PrEP use and worries that conducting screening tests could endanger their safety.

Community-specific barriers included viewing pharmacists as less qualified than sexual health consultants, GPs, or nurses to provide PrEP and a preference for accessing PrEP via GP practices due to concerns of being heard initiating conversations about PrEP in pharmacies. Some individuals preferred GP practices for privacy reasons, while others emphasised the importance of being seen accessing sexual health clinics to combat stigma.

### Motivation facilitators

Motivational facilitators for pharmacists and community members included a preference for independent pharmacy PrEP delivery over other models (e.g., in collaboration with GP, sexual health clinic), believing pharmacies were acceptable places to initiate PrEP, that PrEP use reduces new HIV acquisitions, a perceived personal risk of acquiring HIV and feeling comfortable initiating conversations about PrEP. There was also a belief that accessing PrEP via pharmacies would be more discrete and less stigmatising than from sexual health clinics and potentially address medical mistrust issues among marginalised communities.

Pharmacist-specific facilitators included confidence in conducting sensitive conversations, the belief that community members should initiate conversations about PrEP rather than pharmacists opportunistically identifying people who may benefit from PrEP, anticipation of reimbursement or profit from PrEP delivery, and the potential to enhance professional reputation.

Community member specific facilitators included being confident in pharmacists’ ability to deliver positive STI and HIV results empathically and being able to access PrEP from a pharmacy free of charge.

## Discussion

This is the first research study to explore the barriers to and facilitators of pharmacy PrEP delivery for pharmacists and community members in the UK. The research points to the acceptability and feasibility of pharmacy PrEP delivery in the UK, but demonstrates commonality with barriers and facilitators identified in other countries, particularly the USA (22). Pharmacy PrEP delivery in the UK could offer an important solution to help overcome the current challenges of accessing PrEP via sexual health clinics (e.g., location, opening hours). It could also help to minimise stigma and improve health equity for those underserved by the UKs current model of PrEP provision.

Barriers not previously identified, include community members insufficient knowledge and awareness of pharmacy facilities, the services provided by pharmacists and their roles, qualifications, and capabilities. These barriers have been acknowledged in relation to other pharmacy sexual and reproductive health services (25, 26). However, given the new Pharmacy First initiative enabling patients to be referred into community pharmacy for minor illnesses or an urgent repeat medicine (27), pharmacists expanding roles and increasing responsibilities, particularly since the COVID-19 pandemic, this lack of knowledge among community members is somewhat surprising. Our results therefore highlight the need for further awareness raising about pharmacy facilities (e.g., consultation room), pharmacist’s qualifications and capabilities in addition to the public health services that can be accessed from a pharmacy.

Some barriers identified are suggested to be UK-specific. For example, while pharmacists highlighted a willingness to take on more publicly funded commissioned services, they expressed concern for capacity because of workforce shortages. They also highlighted the potential lack of consistency and subsequent accessibility of service provision between pharmacies due to not all commissioned services being mandatory for all pharmacists to be trained to deliver. This highlights and reiterates previous findings illustrating the current pressures experienced by pharmacists in the UK (28) and could represent significant barriers for the feasibility and sustainability of pharmacy PrEP delivery.

In line with previous research, additional barriers particularly evident among minoritised communities were a concern for confidentiality and stigma (29, 30) due to culture and/or previous negative experiences accessing health care. Although pharmacies were perceived to provide a more discrete less stigmatising environment than sexual health clinics, pharmacists did express unconscious bias and stigmatising attitudes, for example reporting concern that screening for HIV could jeopardize pharmacists’ safety. Results, therefore, highlight the continued need for education and awareness campaigns to help educate healthcare professionals about HIV to reduce institutional, and societal level stigma (26). Additional facilitators to help overcome stigma could include sexual health champions within pharmacies and the co-development of public health services with users to ensure services are designed to accommodate access needs.

Despite the barriers identified, there was a general preference among pharmacists and community members for independent pharmacy PrEP delivery above all other models. Although some community members and pharmacists preferred the idea of collaborative community PrEP delivery with sexual health clinics or GP practices, this preference tended to be associated with a lack of PrEP awareness, including what PrEP delivery would involve, pharmacists’ roles in service provision and the lack of pharmacy facilities to facilitate the screening and monitoring required.

Moreover, numerous solutions to the identified barriers were readily provided. For example, insufficient awareness and knowledge of PrEP in addition to associating PrEP use with different community groups was suggested to be improved by implementing education (e.g., HIV and PrEP) training (e.g., screening, HIV testing) and advertising. Pharmacists and community members also suggested that pharmacy PrEP delivery could be facilitated and more feasible if community members were able to do all the necessary screening tests at home, accessing the pharmacy for the PrEP consultation and dispensing only.

Pharmacists also highlighted that having an appointment system for PrEP delivery (like flu vaccinations), could help alleviate or accommodate the addition of PrEP delivery. An appointment system was also suggested to be facilitative for individuals from minoritised communities, by adding an extra level of privacy for patients. Notwithstanding this, maintaining the ability for patients to ‘walk in’ for PrEP was also recognised as important for improving access and upholding the pharmacy unique selling point.

### Strengths and limitations

Every effort was made to recruit a diverse sample of participants for interviews. Interviewing PrEP users provided valuable insight into their experiences of current PrEP prevision and perceptions of alternative delivery models. Limitations exist in the current research. Due to lifestyle factors, the interviews with sex workers were brief limiting the identification of barriers and facilitators for this group. Further, findings may vary for individuals escorting or working in parlours. Future research should explore the relevance of the current findings to these and other groups who may benefit from improved PrEP access (9).

## Conclusion

The current research, highlights pharmacy PrEP delivery as acceptable to pharmacists and preferred by customers offering an important opportunity to expand PrEP delivery and access. To be feasible, a behaviour change intervention should address barriers and leverage facilitators to support implementation. To increase capabilities and motivation, training and awareness raising for pharmacy staff and community members is needed. To increase opportunities and motivation, environmental and system changes (e.g. facilities, financial reimbursement) are needed.

## Authors contributions

CH, HF, JK, SD, JS, CS, JC, LH, SC, JS and JH contributed to the research planning, CH to data collection and CH and HF to data analysis. CH drafted all versions of the manuscript, and all authors contributed to and approved the final version of the publication.

## Data availability

The data underlying this article are available at data-bris.acrc.bris.ac.uk

