## Supplementary Table 1 for "A qualitative exploration of pharmacists and customers barriers and facilitators to community pharmacy PrEP delivery using the COM-B model of behaviour change"

Barriers of community pharmacy PrEP delivery according to the COM-B with illustrative quotes

|  | **Barriers** | **Illustrative quotes** |
| --- | --- | --- |
| **Capability** |  |  |
| **Psychological** |  |  |
|  | Lack or limited PrEP knowledge e.g., what it is, who it’s for and where it is available from.^TP, BAW, W, YP., pharm^ | “It's (PrEP) either something that you can get the day after you have had unprotected sex, and you suspect it's with someone that’s HIV positive or it's some kind of medication that you take if you have got it (HIV)” W4  “I believe it’s something that can be taken after, if I’m not mistaken and I might be wrong, after sexual intercourse if you believe that you might have contracted it” W2 |
|  | Lack of PrEP awareness including effectiveness of preventing HIV^YP, SW, TP, W, BW. pharm^ | “I’m not aware of it (PrEP), to be honest” Pharm2.  “Some people don’t know about it and that’s the biggest barrier isn’t it” YP3. |
|  | Lack of awareness among the general population ^TP, BAW, YP^ | “I believe in a scale of 10 […] five people know about it” TP11.  “I think people are aware of PrEP but not that much, perhaps I’d give it perhaps a 70% or a 65%” TP18  “Probably out of 10 I think half of it, half” BW14 |
|  | Perceived audience for PrEP (lack of knowledge about all individuals who may benefit from PrEP) ^w,YP, pharm^ | “It’s definitely more the LGBT group that kind of is very geared towards gay men” YP10.  “Quite good for sort of sex workers or people that work in the sex industry” W4.  “most at-risk group of people is young men, I’m presuming, is that fair to say?” Pharm3 |
|  | Lack of and perceived lack of awareness of pharmacy facilities and pharmacist role ^TP,BAW, pharm^ | “They’re just dispensaries” TP7.  “Being a pharmacist, you’re mostly just, well I suppose your day-to-day is writing up prescriptions” YP  “think historically, pharmacies haven’t really necessarily been known to have consultation rooms, although we’ve had to have it for – I don’t know – 10 years or so, so therefore patients aren’t (-). You walk into a GP surgery, and you expect to go into a private consultation room. I don’t think that’s necessarily how all patients perceive pharmacies yet” Pharm7 |
|  | Cold leafletting ^pharm^ | “I think cold bag-stuffing would probably – for some people would be a bit of a resentment when they open the bag at home and just go, ‘Wow. What’s that doing in there?’” pharm10 |
|  | Targeted advertising ^YP, TP^ | “I just think that directly advertising it is a bit invasive” YP3  “That’s going to stigmatise the heck out of it” TP7 |
| **Physical** |  |  |
|  | Pharmacists lacked PrEP training.^YP^ | “Would they have a sort of degree of training in how to deliver that” YP6 |
|  | Lack of skills, training, not feeling competent to deliver PrEP ^pharm^ | “I’m not even remotely feeling that I’m already equipped” Pharm9.  “I feel like when they put extra services on us in pharmacy, they don't give us much training or much equipment or anything and a lot is expected of you so I don't feel comfortable, I don't feel, what's the word, competent?” Pharm6.  “ Most pharmacies don’t have any phlebotomy training so actually taking the blood in a pharmacy at the minute is difficult or will be difficult” Pharm5 |
| **Opportunity** |  |  |
| **Environmental** |  |  |
|  | Lack of skilled staff, staff turnover, inability to hire more staff ^pharm^ | “At the present moment, we can’t get staff, and we can’t get locums, and it’s a real, real challenge” pharm7  “there’s only ever one pharmacists on duty” Pharm3 |
|  | Perceptions of increased pharmacist workload ^pharm, W^ | “Challenges might only be, as I’ve experienced recently, just I think pharmacies are under a lot of pressure and even there you could wait a long time to get booked in or get your prescriptions in and things like that. But I think that’s a widespread issue nowadays” W2.  “If this kicks off then we’re going to be doing this all day and the day-to-day business of checking prescriptions and counselling patients, people at the counter and all the rest of it, would be pushed to the side” pharm10 |
|  | Lack of staff time ^pharm^ | “The only barrier would be time” Pharm1.  “there’s only ever one pharmacists on duty” Pharm3 |
|  | Using pre-existing pathways not appropriate or will not reach all eligible people ^pharm^ | “[…] we’re doing things like morning after pill it’s women who have had sex with a man. So, we’re going to miss out the opportunistic conversations with men who have sex with men or the […] trans or the non-binary population” pharm10. |
|  | Lack of consultation room/space/privacy^W,SW,BAW,YP, TP, pharm^ | “Not just independent pharmacies but the chains as well, they’ve got a converted cupboard that is supposed to be their consultation room […]. A lot of them you struggle to get two people in there” Pharm16.  “It’s private but also, it’s not that private. I think I’ve also seen a curtain one, so it’s almost like a photo-booth kind of thing” YP6. |
|  | Lack of facilities and/or systems ^pharm^ | “Most pharmacies don’t have any phlebotomy training so actually taking the blood in a pharmacy at the minute is difficult or will be difficult” Pharm5.  “I don’t want to be reluctant, but because I cannot see having the facilities for all that, or the staff, I don’t know how viable it is” Pharm2 |
|  | Lack of consistency in service provision/not all staff trained to deliver all services ^YP, pharm^ | “I think the biggest challenge that you sometimes have within pharmacy is that it needs to be consistent i.e. all pharmacies need to be providing it, or none” Pharm7.  “Some have taken the training, and some haven’t done the training.” YP3. |
|  | Pharmacists ability to do all the tests (e.g., bloods)^SW^ | “I would have questions about how they were going to schedule the tests” YP3. |
|  | Lack of medication ^pharm^ | “Another thing is the shortage of medication because pharmacies nowadays we're struggling to get hold certain medication” BAW14. |
|  | Other models of PrEP delivery also considered accessible (e.g., digital PrEP delivery; GP, community setting, sexual health clinic, hospital)^YP,BAW,TP, SW,^ | “From my past injecting lifestyle, I have real trouble to get a vein and it’s also triggering […]. I wouldn’t want to get a blood test there (at the pharmacy)” SW2.  “An online platform where you just fill in the information and you get a discreet delivery, that would be the most ideal way” BAW12. |
| **Social** |  |  |
|  | Preference for GP to do monitoring, taking blood is triggering. ^YP,BAW,TP, SW,^ | “Going to a doctor’s surgery or something like that, then you can sort of see the nurse or someone” SW1  “Feel like it could be done by the GP” YP10 |
| **Motivation** |  |  |
| **Reflective** | Perceived lack of need/decreased risk/lack of interest ^W, YP, SW, pharm^ | “Is there such a great need for it nowadays” Pharm6.  “The problem is I don’t really know because at the moment I don’t feel like I will need it” W2. |
|  | Preference for GP/sexual health delivery ^YP, BAW, W, SW.^ | “I always felt growing up that when you make an appointment with the doctor, it just feels legitimate, and it feels safe, and no one can really say anything. If they see you at the doctor, there is not an immediate suspicion kind of thing, whereas other spaces, it can have a little bit of what’s going on there type of thing and the thing is with the doctor is you can kind of disappear in, no one really knows where you are and it just feels a bit more private I would say and legitimate” W4.  “I’m fine with being seen there (at the sexual health clinic). That’s okay. I want to be seen there” TP8. |
|  | Moral objection to raising awareness of sexual health within community setting^YP^ | “But there is always a bit of a moral objection to having sexual health brought more into the forefront […] So having sex in the pharmacy and discussions about sexual health and sexual practices, even though they probably already have it in pharmacies, but formalising it and bringing in medication for HIV people might find that a little bit […]” YP3. |
|  | Belief that offering PrEP will increase sexually risky behaviours ^pharm^ | “I mean it might make people feel a bit more comfortable in taking risks I guess, so they may feel that they can do anything or do things that they wouldn’t have otherwise done” TP8.  “I mean, it kind of encourages risky sexual behaviour, that's what I feel. Like carry on, do it more” Pharm6 |
|  | Belief that behavioural modification should be promoted above PrEP use ^pharm, BAW^ | “People should change their behaviours to reduce the risk” Pharm5 |
|  | Concern individuals not at need will receive PrEP ^TP pharm^ | “You don’t want a set of people taking it who are not at risk of it” TP7.  “it’s only being supplied to the people that genuinely should be using it” Pharm10 |
|  | Concern about having discussion about PrEP on shop floor ^pharm^ | “Suppose it depends on the individual. Some people are quite open, and quite happy to have a conversation in a pharmacy, in a shop effectively, with other patients. Other people feel more comfortable saying, ‘Can I have a quick word with you in the consultation room?’ So, I think it’s very much on an individual basis, I guess” pharm7  “The downside would be the way in which pharmacies tend to be setup, having that conversation about what you're trying to get, in front of a queue of people, is not necessarily great” YP6 |
|  | If community pharmacy PrEP delivery involved a cost to clients ^YP, TP, BAW, pharm^ | “If you charge for it, that might put people off” TP8.  “I'm just thinking about the cost, because not everybody will be able to afford it” TP20 |
|  | Affordability for pharmacists/lack of compensation ^pharm^ | “I mean, I think it would be an insult to ask us to participate in something and not agree to pay us a sensible amount of money for doing it” pharm9 |
|  | Pharmacies not considered a healthcare source/not considered a space for PrEP delivery. ^YP, W^ | “People want to feel that they have kind of got the proper institutions involved and proper medical professionals, not to say that pharmacists aren’t, but I know what I am like or how I feel” W4. |
|  | Concern about delivering positive HIV result ^pharm^ | “If I were to test somebody for HIV or STI and they become positive and giving that news could be the most challenging thing so far” pharm15 |
|  | Preference to deliver test results by post or online. ^pharm^ | “let them know that their results will follow in 24 hours if they provide their email address […]. I think that would be the safety approach on that” pharm14 |
|  | Concern about pharmacist safety carrying out screening ^pharm^ | “So, for example handling blood samples, especially for people who are potentially HIV positive, they need to know that they should be wearing gloves, they should be making sure there are no needle prick risks. It’s just the basic things around that” BAW5 |
|  | Barriers to initiating conversations on PrEP ^YP^ | “Maybe the women do not feel confident talking about it for cultural reasons” YP1 |
|  | Lack of comfort opportunistically identifying potential customers/initiating conversations about PrEP/concern of causing offense ^pharm^ | “I can imagine if someone came to me and they think they've got an STI with some symptoms and then I'll be like hey, let's talk about HIV, it does scare people” Pharm6. |
| **Automatic** |  |  |
|  | Religion/culture and restrictions on service provision ^pharm^ | “I just think if you went to a Muslim woman who's married and started talking about HIV, they're going to be a bit offended. I think so. I can't imagine them taking it well” Pharm6.  “I’ve had dealings with pharmacists who would refuse to prescribe based on their belief” pharm1 |
|  | Religion/culture and perceived lack of customer trust ^pharm^ | “Their beliefs, their faith. I think there is a lot of mistrust with the establishment I guess” Pharm8. |
|  | Concern being seen accessing PrEP from pharmacy ^BAW^ | “And there’d be a lot of people seeing you [laughs] going to, going to that area and you know, and people looking at you, sort of” BAW5 |
|  | Concern about how pharmacist will treat or judge them.^SW,YP,TP^ | “Girls trying to get the morning after pill in like Boots or in a pharmacy and they’ve just been treated like, you know, there’s been a bit of a hostile attitude towards them. I would never want that to be that manner to be, how someone feels getting PrEP” YP6 |
|  | Stigma and misconceptions about PrEP ^YP, BAW, TP, pharm^ | “If you take PrEP you obviously sleep with a lot of men, you know, it’s kind of that kind of thing” TP10  “Yes, there is a level of stigma that comes with it because a lot of people don’t understand it and they think if you’re taking PrEP you are infected” BAW12 |
|  | Comfort being seen/accessing PrEP from a sexual health service. ^TP, YP, W, BAW^ | “I’m fine with being seen there. That’s okay. I want to be seen there (to show), this is a good thing that other people should also do and also, I’m a safe person” TP8 |
|  | PrEP use associated with risky behaviours ^pharm^ | “I mean, they’re potentially going to get infected by engaging in that activity and obviously, if they’re thinking about using PrEP then they know that that’s a risk, so they know it’s a risk otherwise they wouldn’t come in, or they wouldn’t be looking to get this medication, so the patient themselves will understand that there’s a risk in what they’re doing but they’re going to do it” pharm3 |

TP=trans person, YP=Young person, BAW= Black African woman, W=Woman, SW=Sex worker, pharm=pharmacist
