## Supplementary Table 2 for "A qualitative exploration of pharmacists and customers barriers and facilitators to community pharmacy PrEP delivery using the COM-B model of behaviour change"

Facilitators of community pharmacy PrEP delivery according to the COM-B model with illustrative quotes

|  | **Facilitators** | **Illustrative quotes** |
| --- | --- | --- |
| **Capability** |  |  |
| **Psychological** |  |  |
|  | Continued education/training for HIV/PrEP  ^pharm^ | “[…] feeling confident with all the details, the doses, and everything. We need the training in order to start even doing the conversations” Pharm2.  “Training, to make sure I know what I’m talking about” Pharm12. |
|  | Awareness/improving awareness of PrEP including effectiveness of preventing HIV ^YP, TP, BAW, pharm^ | “Spreading the word about PrEP” TP10  “I think the first thing is to make people aware”. Pharm11  “it would be good to raise awareness and some way of people, letting people know that it's available” Pharm8  “maybe if awareness was increased first, then people would think about it more” Pharm6 |
|  | Good knowledge of clients/being able to identify potential clients. ^pharm^ | “I would have said that it was open to anybody who felt they themselves could be at risk from their sexual practices” Pharm9  “In terms of who can use it, I understand that it is most people that can use” Pharm5 |
|  | Improving awareness of pharmacy facilities and roles of pharmacist for public health service provision including PrEP ^YP, W^ | “Getting people to know that it’s in pharmacies” YP3  “then they need to be making it known that they could go to a pharmacy and be given counselling and advice” Pharm9 |
|  | Advertising PrEP through a variety of different platforms (e.g., socials, health settings, education, community)/advertising in general ^YP, TP, BAW, W, SW, pharm^ | “If I am in a waiting room and I am waiting around, I often just see what leaflets they have around or what’s on the walls and take note of them”. W4  “Always an opportunity now so Facebook, Instagram, whatever. These are all ways where you can spread the word amongst communities using or accessing those things” Pharm10 |
|  | Targeted advertising  ^pharm^ | “so, they’d probably be better targeted in terms of getting the message out that this is available” Pharm3 |
| **Physical** |  |  |
|  | Knowing PrEP was available form pharmacies ^YP, pharm^ | “Getting people to know that it’s in pharmacies” YP3 |
|  | Knowing PrEP access will be kept confidential ^YP, TP, BAW^ | “It (accessing PrEP) was confidential, and it shouldn’t be discussed with my GP” BAW5  “As I understand sexual health clinics do not reveal your personal stuff to your doctor, to your GP, so I think also I’d be concerned if the pharmacies would also do the same” TP18 |
|  | Knowing pharmacists had specific training/education for PrEP and provision was certified ^TP^ | “I’d want to be sure if it’s certified and if the personnel are qualified” TP18 |
|  | Pharmacists being educated to be prescribers ^pharm^ | “Pharmacists will be able to prescribe medications to patients rather than them going to the GP” Pharm5. |
|  | Having PrEP expert on call for advice ^pharm^ | “Someone who’s experienced in it, whether it is a sexual health clinic or whatever, somebody who would be happy to take calls from a pharmacy” Pharm16. |
|  | PrEP delivered via a PGD ^pharm^ | “Pharmacists won’t have any issue with that delivering PrEP as a service on the PGD” Pharm16. |
|  | Additional training including how to have sensitive conversations. ^pharm^ | “But obviously everyone would have to be trained up to deliver that to the level that they needed to” Pharm10 |
|  | Knowing that pharmacists were able to do screening. ^YP, TP, W,^ | “It would be okay as long as I knew that they had all the tools” W2 |
|  | Prior experience of doing screening tests ^pharm^ | “We already do – or have done over the years – blood and – cholesterol and glucose testing. So, finger-pricking patients in a sensible environment, it’s doable”. Pharm10 |
| **Opportunity** |  |  |
| **Environmental** |  |  |
|  | AI to alleviate dispensing responsibility. ^pharm^ | “One of the pharmacies that I work in, we have this advanced dispensing system called Titan” BAW15 |
|  | Other staff can take some of the workload. ^pharm^ | “I think going forward more of this is going to happen anyway, I think as more services are coming in for us we’ll be in the back and the staff at the front will have to take on more roles” Pharm1.  “we’re trying to build our team up with other qualifications so they can take some of the workload away from the pharmacist to be able to spend more time with patients” Pharm3 |
|  | Inaccessibility of sexual health clinic PrEP provision (e.g., inconvenient location of sexual health clinic, waiting lists) ^TP, SW, TP, YP^ | “There’s obviously physical distance barriers like I live 45 minutes away from sexual health clinic” YP10.  ‘With a chaotic lifestyle of using and […] keeping appointments is quite hard and […] going to a sexual health clinic, that’s not easy’ SW2. |
|  | Pharmacy’s convenient location, accessibility, opening hours and/or quick service for PrEP delivery including screening ^YP, TP, BAW, SW, W, pharm^ | “I think that would be much easier. I live one minute away from a pharmacy. Pharmacy is a very local thing so that would make it a lot easier for people to get PrEP” YP10.  “It would be a lot easier for people to come to us rather than driving into central (name of area) every time” Pharm5 |
|  | Appointment system ^BAW, YP, W, pharm^ | “In terms of the screening though, maybe I would like for the pharmacist to be aware that I was going and maybe meet me” W2  “More like an appointment session really book an appointment with the pharmacist”. BAW15 |
|  | Walk in service ^YP, pharm^ | “I cannot hang on the telephone to make an appointment and, no, it’s just go there” YP1 |
|  | Use of existing pathways (e.g., c-card, drug substitution) ^SW, TP, pharm^ | “It would be really good if when I access the C-Card service and got condoms they then also took the opportunity to go hey, do you know we do these other services? That would be the ideal time” TP8 |
|  | Private consultation rooms/private space ^pharm^ | “they’re all supposed to be soundproof so they all should be private and secure in that respect” Pharm10 |
|  | Having facilities to do screening and monitoring ^pharm^ | “We’d have to have the correct facilities to dispose of biological waste and all those sorts of things” Pharm12  “I mean as long as we have the equipment and be able to send the monitoring back to you,” Pharm8. |
|  | Easier access, less bureaucracy ^TP, BAW, YP, W, SW^ | “Yeah so, I would not be going through a lot of protocol. I would just get the pharmacist and then we will not have much pressure” YP17. |
|  | Shorter waiting lists/quick service ^pharm^ | “We don't have the waiting times that a GP has or a sexual health clinic because they can just put anything on us whenever they want, basically” Pharm6. |
|  | Consistency in service provision ^pharm^ | “Every pharmacist knows what that service is, and every pharmacist can deliver it” Pharm4 |
|  | Acceptability of and/or preference to do home testing ^TP, BAW, YP, SW,pharm^ | “So, I think it would come down to having a list of sort of approved, HIV testing kits and all the organisations that provide them and their sort of range of accuracy. And if one presented with that […] one going, ‘Oh, I had this test done by” YP10.  “I think in the pharmacy setting somebody should come with a pre agreed upon acceptable test that you just show to the attendant, they know it is the standard one and it must be legit, they just give you your medication” BAW12. |
|  | Preference for screening to be done by someone else instead of at home for reliability ^TP, YP, BAW,^ | “I think I’d rather have someone else do it for me and rather there’d be that personal element ‘cause I’d feel like I was doing it wrong or I wasn’t doing” YP10. |
|  | Receiving ongoing monitoring at a pharmacy ^BAW, TP^ | “I think at […] a pharmacy” TP18 |
|  | Collaborative PrEP delivery ^TP,YP, pharm^ | “I think a combined service” TP6  “I think that all these tests could happen in the clinic, and then the pharmacist could be a follow-up contact point, probably” Pharm2 |
|  | Collaborative service for monitoring - Clear referral pathway ^pharm^ | We could certainly do the follow-up conversation but if that involved having kidney testing for example then we’d have to then have a referral pathway into a – either their local GP practice or – does the sexual health clinic request those diagnostic testing sort of thing test?” YP10. |
|  | Independent service provision ^BAW, pharm^ | “Making the PrEP available, the testing, the screening, everything available from the pharmacy would work far better” BAW14  “I would love that we were able to counsel, advise, and supply, yes […]. I think the role of pharmacy has so much more than merely providing tablets” Pharm9. |
| **Motivation** |  |  |
| **Reflective** |  |  |
|  | Perceived importance of PrEP, moral imperative and belief that PrEP would reduce HIV acquisitions ^YP, TP, BAW,SW, W, pharm^ | “I think it’s very important. I perfectly valid service that is necessary” Pharm16. |
|  | Preference for only prescribing/dispensing from pharmacy ^YP, BAW, TP,^ | “If there was a process involved that I was sent home a test kit that went to the lab and then a prescription and I could get more PrEP, then I’d be very happy with that. And so I would go, I would only go to the pharmacy kind of thing” TP6 |
|  | Preference for pharmacy delivery, Belief that pharmacy is an acceptable place to initiate and receive PrEP ^YP, pharm^ ^BAW, YP, SW, TP, pharm^ | “I would say the pharmacy would be the first option” YP1.  “Making the PrEP available, the testing, the screening, everything available from the pharmacy would work far better” BAW14 |
|  | Belief pharmacy delivery would improve access and reduce HIV acquisitions ^BAW,^ | “It’s going to be easier that way and it’s going to help prevent a lot of people from getting infected with HIV” BAW13. |
|  | Perceiving customers to be comfortable accessing PrEP from a pharmacy. ^pharm^ | “Think ‘they’d feel great. ‘They’d want to […] come to the pharmacy” Pharm6. |
|  | Previous use of pharmacies ^YP^ | “I’ve gone to any community health service before well I would just feel okay to be honest for the fact that I said what I’m looking for and then they try to give me a good service” TP17 |
|  | Assistance with costs/PrEP provision was free ^TP,pharm^ | “If it could still be free in a pharmacy and making it mandatory to order from a pharmacy to make sure it's free, then I think for us it will be more preferred” TP20 |
|  | Costs reimbursed/PrEP seen as a source of profit ^pharm^ | “I think it's all down to income, […] I think all pharmacists nowadays are very keen on services, because it's the only way we can get some income. So, as long as it’s funded, ’ can't see any issue” Pharm14 |
|  | Belief PrEP delivery would add positively to reputation/career development. ^pharm^ | “So, the more services we offer, then the better our profession is perceived by the patients” Pharm7 |
|  | Recognition of pharmacists as health care providers/well qualified/aligned with scope of work ^pharm^ | “So, I think especially now a lot more like with the consultations coming over to the pharmacist and whatever, I think pharmacy staff and pharmacists are a lot more -à have a broader range. So I don’t think it would be an issue at all and certainly the pharmacists I know that would be the most sensible place for it to be issued” Pharm16 |
|  | Comfortable providing information and having sensitive conversations with clients ^pharm^ | “it’s something […] that I’ve done and certainly I’m fairly comfortable about doing it” Pharm9 |
|  | Comfortable seeking a pharmacist for PrEP ^BAW^ | “Pharmacy then I think I’ll feel more comfortable” BAW15 |
|  | Comfort/confidence in pharmacists’ professionalism to deliver positive result ^YP, TP, BAW, W, SW, pha’m^ | “I'm guessing if so’eone's trained to break that sort of news, that they’re someone who is capable of that doing” TP7 |
|  | Preference to receive test results at home ^YP, TP^ | “I’d probably just prefer to get it on my own because that would be quicker and easier and also just, I don’t want be processing that in the middle of the pharmacy” TP8 |
|  | Need for signposting after positive result ^TP, YP^ | “To be signposted to the right place for more information” TP18 |
|  | Ensuing pharmacist safety doing screening tests ^pharm^ | “One of the fundamental points is the safety of the service, because we have to protect the pharmacists from catching any– because he was delivering the service, catching a virus, ‘that’s a very important thing” Pharm14 |
|  | Need for positive results to be delivered empathically ^TP, BAW, YP,^ | “Knowing that he will sympathise you know” BAW5  “I think I’ll really need to have somebody to support me at that time and guide me” TP18 |
|  | Comfortable being approached about PrEP ^TP, TP, BAW, YP, W^ | “Even if it’s in front of people in the presence of loads of people, I think I’d be comfortable”” TP18. |
|  | Comfort opportunistically identifying potential customers. ^pharm^ | “I feel fine. Well, from my side it’s like one of the normal services I provide. ’ can't see any issue on that” Pharm14. |
|  | Belief that customers should approach pharmacists for PrEP ^pharm^ | “If it was something that was raised by the person, then we could kind of go into i’, it's not something that we would I suppose randomly start to discuss with people” Pharm8 |
| **Automatic** |  |  |
|  | Lack of trust in medical professionals GP/level of confidentiality/stigma ^BAW,YP,TP, SW^ | “The problem is if you are a medical professional, and you see these records on somebody’s file you know exactly what they relate to. So, it is not really that confidential” BAW12  “Black people, don’t really have trust in the doctors and the system a bit, I think people feel neglected in a way or looked down upon” TP18 |
|  | Pharmacist customer relationship ^SW, pharm^ | “Somewhere that they are familiar, have that conversation in a familiar environment with someone that they know, that would probably help a lot” TP8.  “You are also very good at building trust and carrying people along. And the data supports that the majority of the population find talking in a pharmacy is very comfortable” Pharm11 |
|  | Belief that pharmacy delivery would help with stigma. ^YP, TP, BAW, W, SW, pharm^ | “Because you’re just going to the pharmacy. There’s a lot of reasons to go to the pharmacy” TP8  “I think because it could make it easier and it’s always very private, I think it could help a lot. It would also maybe normalise the process for someone who has been led to think that they are different because they need it or they have done something wrong because they need it” W2 |
|  | Stigma- Feeling uncomfortable accessing sexual health services. ^YP, TP, BAW, W, SW, pharm^ | “Accessing a sexual health clinic for some people can create serious problems for them. If someone from a religiously conservative family or community is seen walking into a sexual health clinic before they are married, that could be big problems for them” W4. |
|  | Belief that pharmacy less stigmatising and more discrete ^TP, pharm^ | “I think that would also be great because I feel it will be discreet in a way because, you know, going to a pharmacy you can be going to pick anything, not necessarily something related to sexual health or, yeah, so I think it’s a nice idea too” TP18. |

TP=trans person, YP=Young person, BAW= Black African woman, W=Woman, SW=Sex worker, pharm=pharmacist
